# Left ventricular systolic dysfunction in long-term aficamten therapy for obstructive HCM is rare, transient, and well-tolerated

**DOI:** 10.64898/2026.11.23.26360829

**Authors:** Daniele Massera, Anjali Owens, Theodore P. Abraham, Roberto Barriales-Villa, Robert M. Cooper, Perry M. Elliott, Martin S. Maron, Michael E. Nassif, Artur Oreziak, Sara Saberi, Scott D. Solomon, Albree Tower-Rader, Justin Godown, Stephen B. Heitner, Daniel L. Jacoby, Stuart Kupfer, Xueli Liu, Fady I. Malik, Regina Sohn, Jenny Wei, Ahmad Masri

## Abstract

**Aims:** Aficamten, a novel cardiac myosin inhibitor recently approved in several regions for adults with symptomatic obstructive hypertrophic cardiomyopathy (oHCM), requires structured echocardiographic monitoring to assess for left ventricular (LV) systolic dysfunction (LVSD). We aimed to describe the incidence, severity, trajectory, and clinical correlates of LVSD during aficamten therapy in patients enrolled in FOREST-HCM.

**Methods:** Serial echocardiographic examinations were performed to inform investigator-driven dose adjustments and safety monitoring. Patients were categorized by on-treatment LV ejection fraction (LVEF): any incidence of LVSD (<50%) or no LVSD (always ≥50%). A prespecified subgroup analysis evaluated patients with any LVEF ≥50% to <55%.

**Results:** Among 410 patients (mean [SD] age 60.2 [12.8] years, 43.7% female), 12 (2.9%; exposure adjusted incidence rate [EAIR] 2.1/100 patient-years) developed LVSD (none with LVEF <40%); only one event was corroborated by the core laboratory. LVEF recovered to ≥50% in 11 (91.7%) patients; 10 following dose reduction and one after transient interruption. One patient had persistent LVSD, having also experienced LVSD while receiving placebo in the parent study. Concurrent non-serious clinical heart failure occurred in two patients (0.5%; EAIR 0.3/100 patient-years), resolving in one, while the other (with prior LVSD on placebo) permanently discontinued aficamten. Additionally, 47 patients (11.5%; EAIR 8.8/100 patient-years) demonstrated LVEF ≥50% to <55% and remained on aficamten without dose reduction or associated adverse events.

**Conclusion:** LVSD was infrequent, mild, reversible, and rarely associated with transient non-serious heart failure symptoms. LVEF ≥50% to <55% appeared clinically benign, supporting structured echocardiographic monitoring and individualized dose optimization.

**Graphical Abstract:** 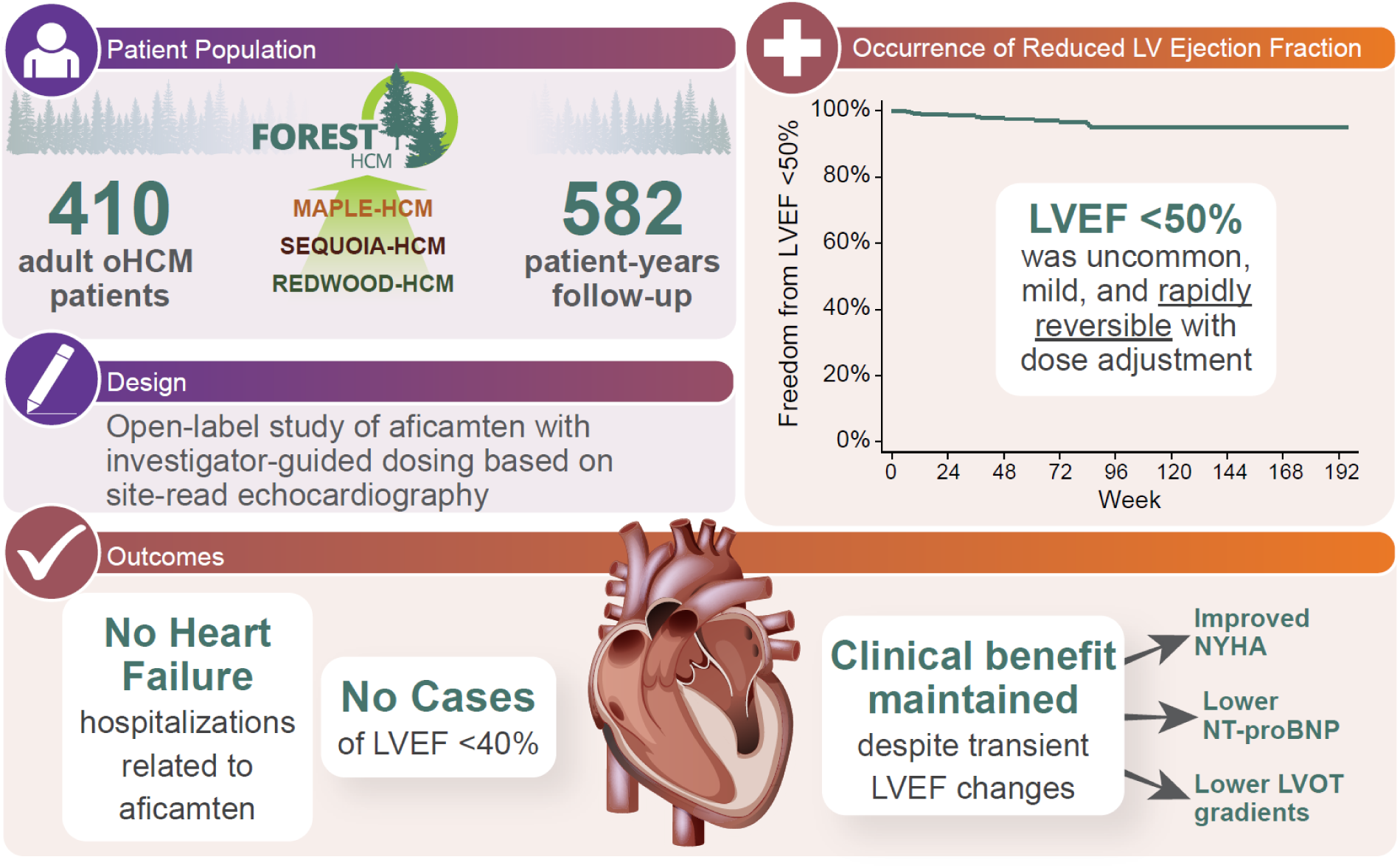

**Trial registration number:** NCT04848506

## Introduction

Obstructive hypertrophic cardiomyopathy (oHCM) is characterized by left ventricular hypertrophy, hypercontractility and systolic anterior motion of the mitral valve, which drives the pathophysiology of left ventricular outflow tract obstruction (LVOTO) and is the primary aetiology of exercise intolerance and heart failure symptoms.^1^ Cardiac myosin inhibitors (CMI) alleviate LVOTO by targeting myocardial hypercontractility through a decrease in actin-myosin cross-bridging, thereby improving symptoms, exercise capacity, and quality of life.^2,3^ However, occasionally, an excessive reduction in left ventricular ejection fraction (LVEF) may be observed and is an expected pharmacodynamic consequence of the mechanism of action. The impact of low LVEF excursions on overall safety, their reversibility, and clinical significance remains incompletely explored.^4^

Experience with CMIs has underscored the importance of systematic echocardiographic monitoring. The incidence of left ventricular systolic dysfunction (LVSD) in mavacamten-treated patients with oHCM ranged between 5% and 14% in placebo-controlled clinical trials.^2,3,5^ By comparison, the incidence of LVSD in oHCM clinical trials of aficamten, a next-in-class cardiac myosin inhibitor with unique pharmacokinetic and pharmacodynamic characteristics, was numerically lower, with an incidence of 3.5% (vs. 0.7% with placebo) in SEQUOIA-HCM (Safety, Efficacy, and Quantitative Understanding of Obstruction Impact of Aficamten in HCM; NCT05186818), 1% in MAPLE-HCM (Metoprolol vs Aficamten in Patients With LVOT Obstruction on Exercise Capacity in HCM; NCT05767346) and 3.4% in the long-term open-label study, FOREST-HCM (Follow-Up, Open-Label, Research Evaluation of Sustained Treatment With Aficamten in HCM; NCT04848506).^2,6,7^ Aficamten studies utilize a dose-adjustment regimen based on site-read LVEF, where after dose titration is complete, aficamten doses are maintained if LVEF is ≥50 to <55%, reduced if ≥40% but <50%, and temporarily held if <40%.^8,9^

FOREST-HCM is an open-label study designed to evaluate the long-term safety and efficacy of aficamten in a larger cohort of patients with symptomatic oHCM using a flexible titration and monitoring paradigm that mirrors the approved labelling of aficamten.^7^ The study incorporates echocardiography-guided dose adjustments and allows two potential monitoring strategies for LVEF assessment (every 3 or every 6 months) once target doses are achieved. Consistent with anticipated real-world clinical practice, dosing adjustments and the frequency of monitoring are determined by the treating investigator. This approach aligns with the approved prescribing framework for aficamten, which emphasizes routine echocardiographic monitoring of LVEF prior to treatment initiation, during dose titration, and periodically during maintenance therapy. These safeguards are implemented through a Risk Evaluation and Mitigation Strategy (REMS) programme that requires prescriber certification, patient enrolment, and documentation of serial LVEF assessments to support ongoing treatment.^10^

In the present analysis, we sought to explore the trajectory of LVSD in aficamten-treated patients with oHCM to quantify the incidence and clinical significance of LVEF <50%, as determined by the treating investigator. Importantly, this analysis focuses on site-read echocardiographic assessments that directly informed treatment decisions, rather than retrospective core laboratory adjudication. As such, these findings provide a clinically relevant evaluation of how aficamten is managed in practice and offer insight into the performance of the approved monitoring strategy in a real-world setting. We aimed to characterize the long-term safety profile of aficamten, including the reversibility of systolic dysfunction following dose adjustment. We also evaluated associations between LVEF changes and long-term efficacy outcomes including functional capacity, quality of life, LVOT gradients, and the key clinical events of heart failure, atrial fibrillation, and mortality.

## Methods

### FOREST-HCM study design and population

The FOREST-HCM population for this analysis includes adults (≥18 years of age) with oHCM and LVEF ≥55% who have previously completed a parent aficamten study (REDWOOD-HCM, SEQUOIA-HCM or MAPLE-HCM).^2,9,11^ The study protocol was approved by the institutional review board or ethics committee at each participating site, and all patients provided written informed consent prior to enrolment. The study was conducted in accordance with the principles of the Declaration of Helsinki and Good Clinical Practice guidelines.

Dose titration and safety monitoring were guided by site-read echocardiograms. Patients were excluded from participating in FOREST-HCM for prior history of LVEF <45% or stress cardiomyopathy and for LVEF <40% while being treated with aficamten during a parent study leading to treatment interruption. No patient in SEQUOIA-HCM or MAPLE-HCM met the criterion of confirmed LVEF <40% leading to aficamten treatment interruption.

The study schema for FOREST-HCM is presented in **Supplemental Figure 1**.

### Treatment and monitoring

Once-daily oral aficamten was initiated at 5 mg in all patients and titrated (in 5 mg increments) based on site-read echocardiograms to a maximum dose of 20 mg to achieve a Valsalva LVOT gradient <30 mmHg while maintaining LVEF ≥50%. Echocardiograms were performed approximately two weeks after any dose adjustment. After the target dose was achieved, monitoring echocardiograms were conducted every 12 weeks for patients with LVEF ≥50% to <55% and every 24 weeks for those with LVEF ≥55%, or sooner if clinically indicated. Dose titration was at the discretion of the site investigator rather than protocol-mandated, allowing incorporation of clinical judgement. Aficamten dose was reduced by 5 mg for LVEF ≥40% to <50% and temporarily interrupted for LVEF <40%. Additionally, echocardiograms were interpreted by the Cardiovascular Imaging Core Laboratory (Brigham and Women’s Hospital, Boston, MA, USA) for comprehensive assessment. However, these data were not used for dose titration.^12^

### Subgroup definitions

Patients were categorized into two groups based on LVEF during treatment: 1) any LVSD (LVEF <50%) detected and 2) LVEF always ≥ 50%. We further examined a pre-specified subgroup of patients with any instance of LVEF ≥50% to <55%.

### Outcomes

The main objective of this analysis was the safety of aficamten in each subgroup with a particular focus on dose adjustments, treatment interruptions, and clinical events (heart failure, atrial fibrillation, and mortality) related to occurrences of LVSD. Secondary endpoints included changes in additional parameters that were assessed at each study visit, including LVEF, New York Heart Association (NYHA) functional class, Kansas City Cardiomyopathy Questionnaire-Clinical Summary Score (KCCQ-CSS), resting and Valsalva LVOT gradient, N-terminal pro–B-type natriuretic peptide (NT-proBNP), and high-sensitivity cardiac troponin I (hs-cTnI).

### Statistical analysis

Baseline characteristics, echocardiographic parameters, and clinical outcomes were summarized according to the specified groups using descriptive statistics. Continuous variables are presented as means and standard deviation or medians and interquartile range, as appropriate.

Categorical variables are presented as counts and percentages. Given the low frequency of systolic dysfunction, comparisons between groups were exploratory and descriptive in nature, and no formal hypothesis testing was prespecified for this analysis. All analyses were conducted using observed data, and there was no imputation for missing data. Analyses were performed using SAS Enterprise Guide version 8.3.

## Results

### Baseline characteristics

A total of 410 patients with oHCM, mean (SD) age 60.2 ±12.8 years and 43.7% female, enrolled in FOREST-HCM from May 28, 2021, to May 9, 2025, were included in this analysis (**Graphical Abstract**). Mean (SD) baseline LVEF was 68 ± 6%, resting and Valsalva LVOT gradients were 55 ± 37 mm Hg and 93 ± 44 mm Hg, respectively. At baseline, the majority (N=396, 96.6%) of patients were symptomatic with 59.5% (N=244) classified as NYHA class II and 37.1% (N=152) NYHA class III, with a mean KCCQ-CSS score of 70.3 ± 18.9. There were 12 patients (2.9%), who had at least one occurrence of a site-read LVEF <50% (exposure adjusted incidence rate [EAIR] 2.1 per 100 patient-years) and none with LVEF <40%. Only one of the occurrences of LVEF <50% among these 12 patients was corroborated by the core laboratory. Patients with and without LVEF <50% were broadly similar with respect to age, sex, race/ethnicity, baseline symptom burden (KCCQ-CSS and NYHA class), background medical therapy, and biomarkers (NT-proBNP and hs-cTnI). Patients with LVEF <50% had lower mean (SD) LVEF at baseline (63.8% ± 4.7 vs. 68.3% ± 5.7, p=0.007), lower Valsalva LVOT gradients (70.8 ± 14.6 mm Hg vs. 93.3 ± 44 mm Hg, *p*<0.001), longer QRS durations (128.7 ± 34.3 ms vs. 105.7 ± 20.9 ms, p=0.041), and longer QTc intervals (466.7 ± 38.8 ms vs. 444.7 ± 30.3 ms, *p*=0.015) compared with those without LVEF <50% (**Table 1**). Cumulative exposure was 582 patient-years, and the median follow-up was 96.2 weeks for those with LVSD and 79.4 weeks for those without.

**Table 1.**
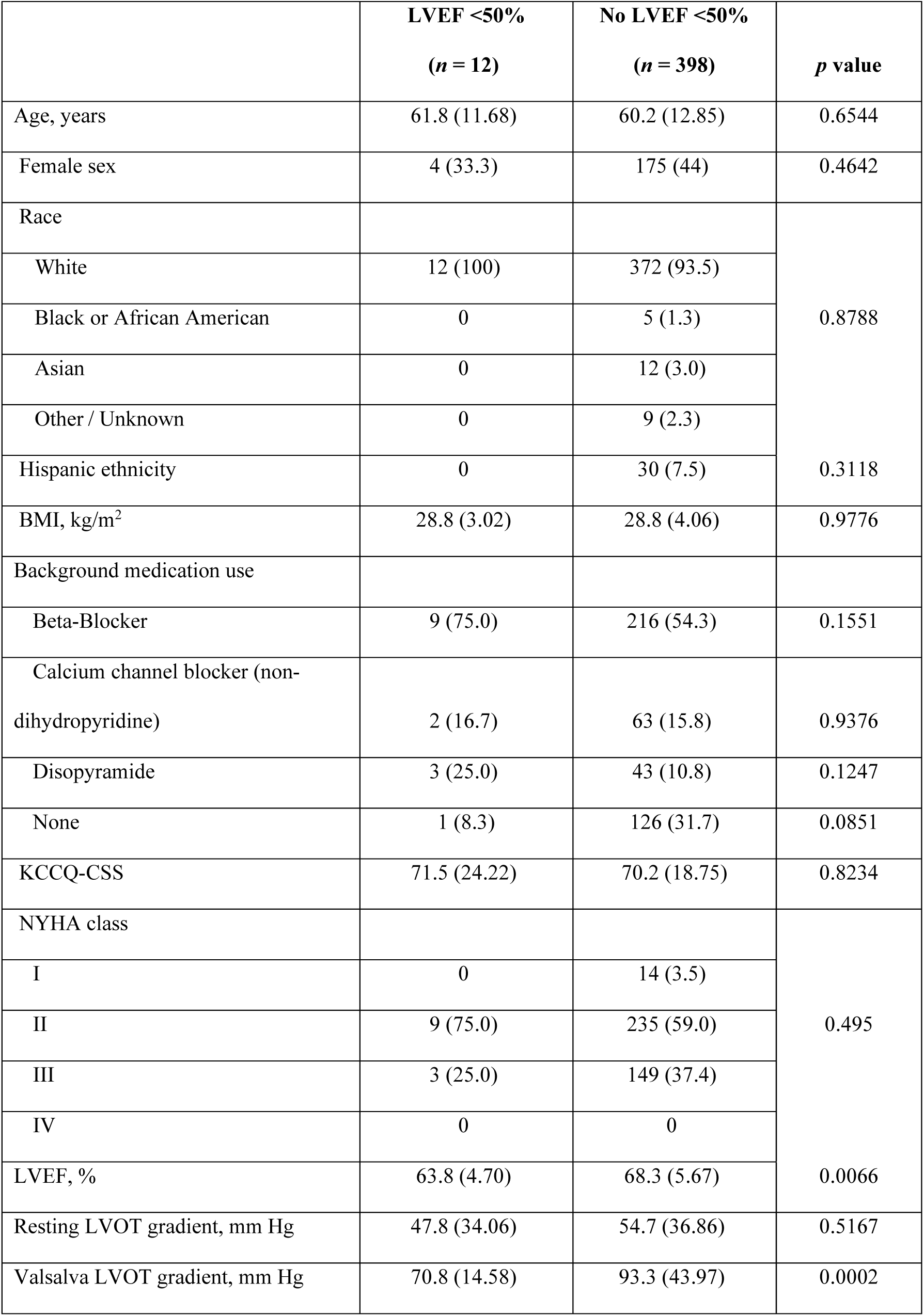
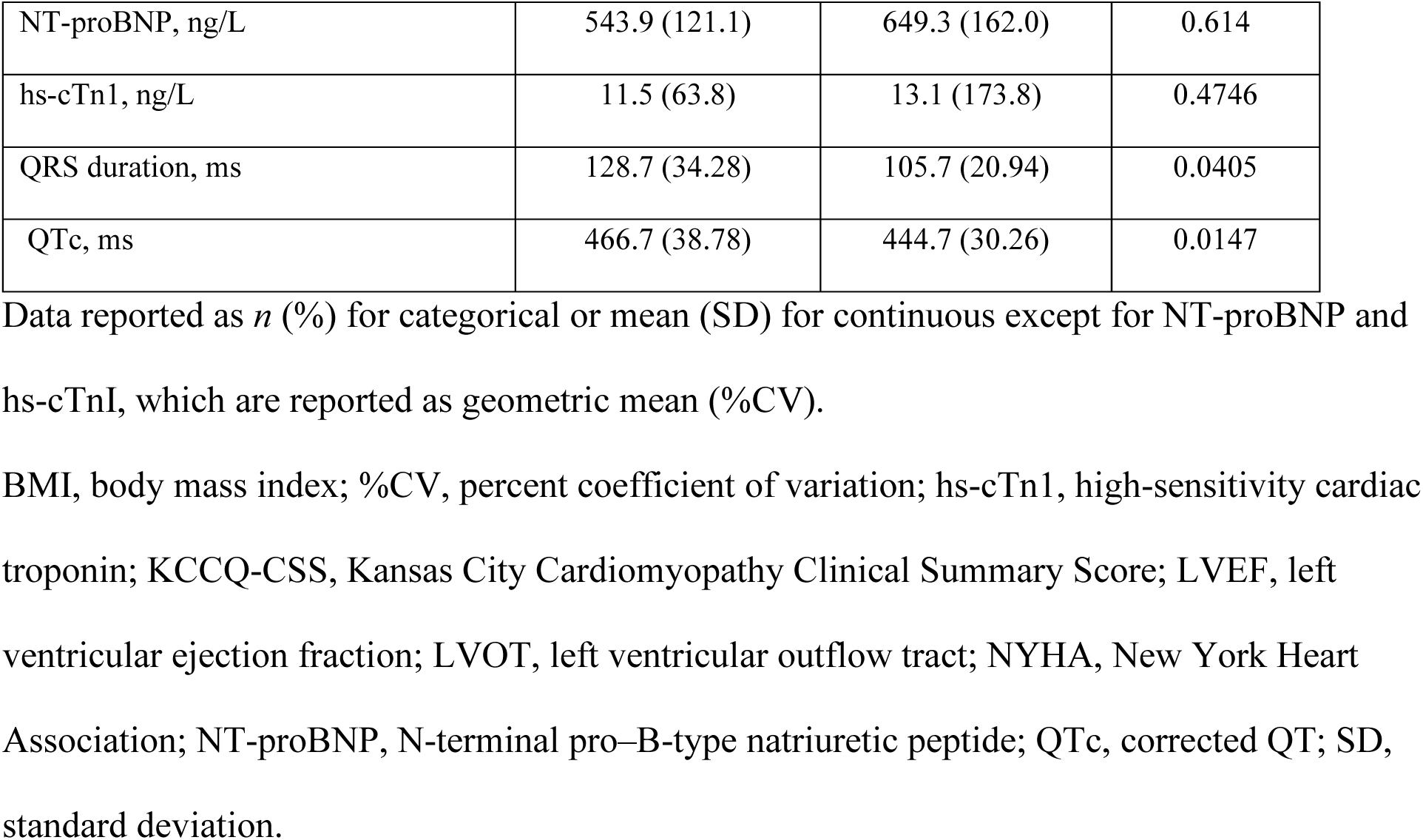
Demographics.

### Dosage

The last recorded dose of aficamten in patients was 5 mg in 35 (9%), 10 mg in 80 (20%), 15 mg in 82 (20%), and 20 mg in 205 (51%) patients. Maintenance doses of aficamten for each group are presented in Supplemental Table 1. There was no difference in the mean maintenance dose in the LVEF <50% group (12.9 mg ± 4.5) and the LVEF ≥50% group (15.1mg ± 4.7) (p=0.114). The doses of aficamten coincident with the initial detection of LVSD were: 10 mg in 1 (8.3%), 15 mg in 7 (58.3%) and 20 mg in 4 (33.3%) patients. No patients were on 5 mg at the time of initial detection of LVEF <50%.

### Trajectory of patients with LVSD

In the 410 patients, 3764 post-baseline echocardiograms were completed. Among the 12 patients with at least one occurrence of LVSD, there were a total of 21 (0.6% of post-baseline echocardiograms) LVSD episodes. Median time to the first LVEF <50% was 37 (IQR 11 to 67) weeks after initiation of treatment and no participant had an LVEF <40%. Baseline characteristics and LVSD event profiles of each participant with LVSD are described in **Table 2**. All patients demonstrated improvement in NT-proBNP relative to baseline at the time of LVSD detection, with no worsening in NYHA functional class at the time of the lowest recorded LVEF. Among the 10 patients with KCCQ-CSS assessed at the time of the low LVEF event, none experienced a clinically meaningful decline. Additionally, Valsalva LVOT-G remained ≤30 mm Hg in 10 of the 12 patients at the time of LVSD (**Table 2**). After detection of low LVEF excursions, all 12 patients underwent per protocol dose reduction with subsequent increase in LVEF to ≥50% in 11 (91.7%) patients; 10 after dose reduction alone and 1 following temporary interruption. Median time to recovery was 2.1 weeks (IQR 2.1 to 4.1). All 11 patients who recovered LVEF following dose reduction were able to continue aficamten. One participant permanently discontinued treatment due to recurrent LVSD following initial recovery of LVEF to >50% after dose reduction; however, this participant was also noted to have had LVSD while receiving placebo during the parent study. **Figure 1** illustrates the trajectory of LVEF in all patients with LVEF <50% and **Figure 2** highlights the overall freedom from LVSD. In patients with LVSD, the mean LVEF was 45.1% ±2.5 at the time of detection and increased to a mean of 51.7% ±5.9 at the next visit, after per protocol dose reductions (**Table 3**).

**Figure 1.**
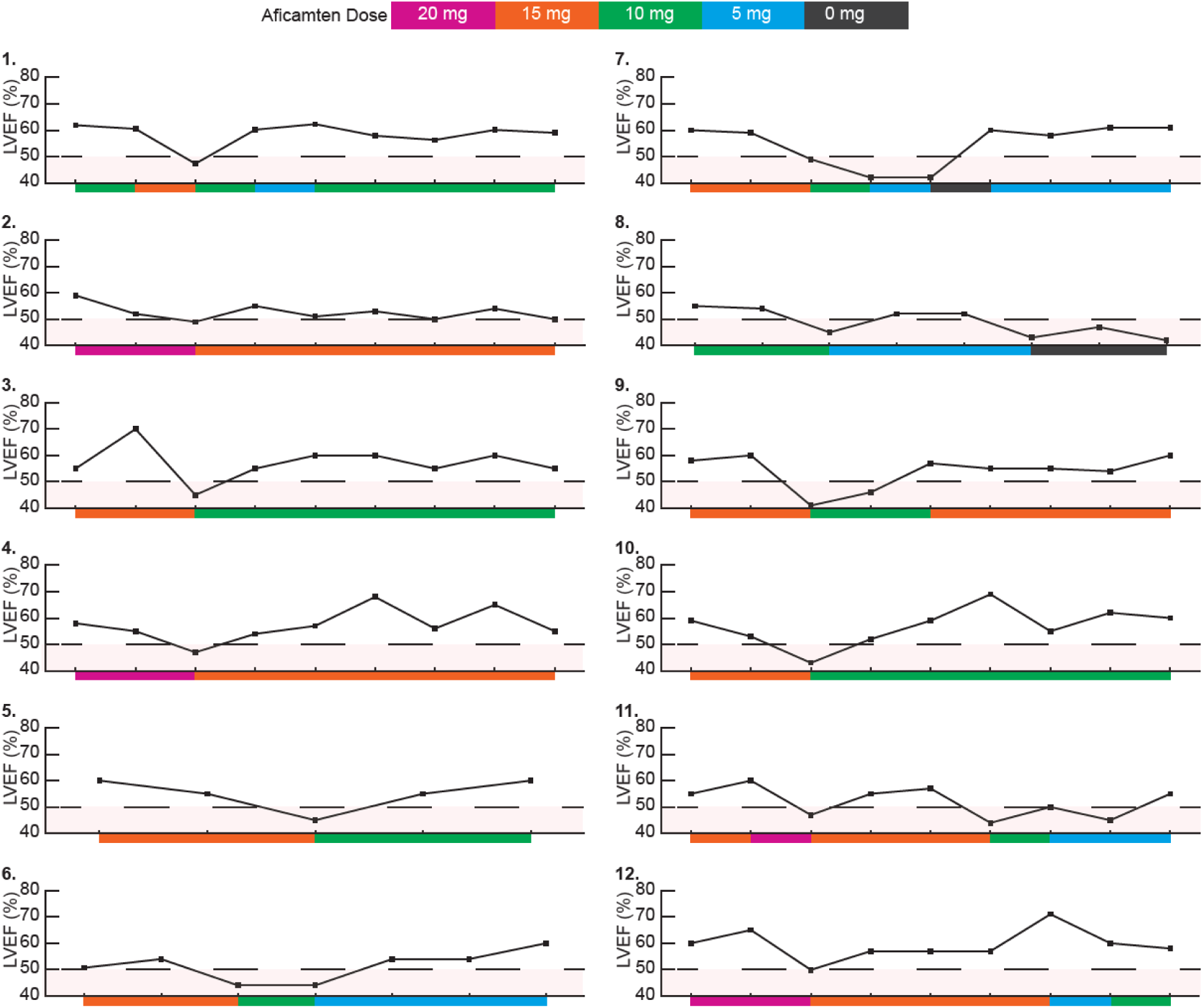
Trajectory of LVEF in patients with at least one LVEF <50%. For each patient, the two data points immediately preceding the occurrence of LVEF <50% are shown to illustrate antecedent LVEF trends prior to the low EF event. EF, ejection fraction; LVEF, left ventricular ejection fraction.

**Figure 2.**
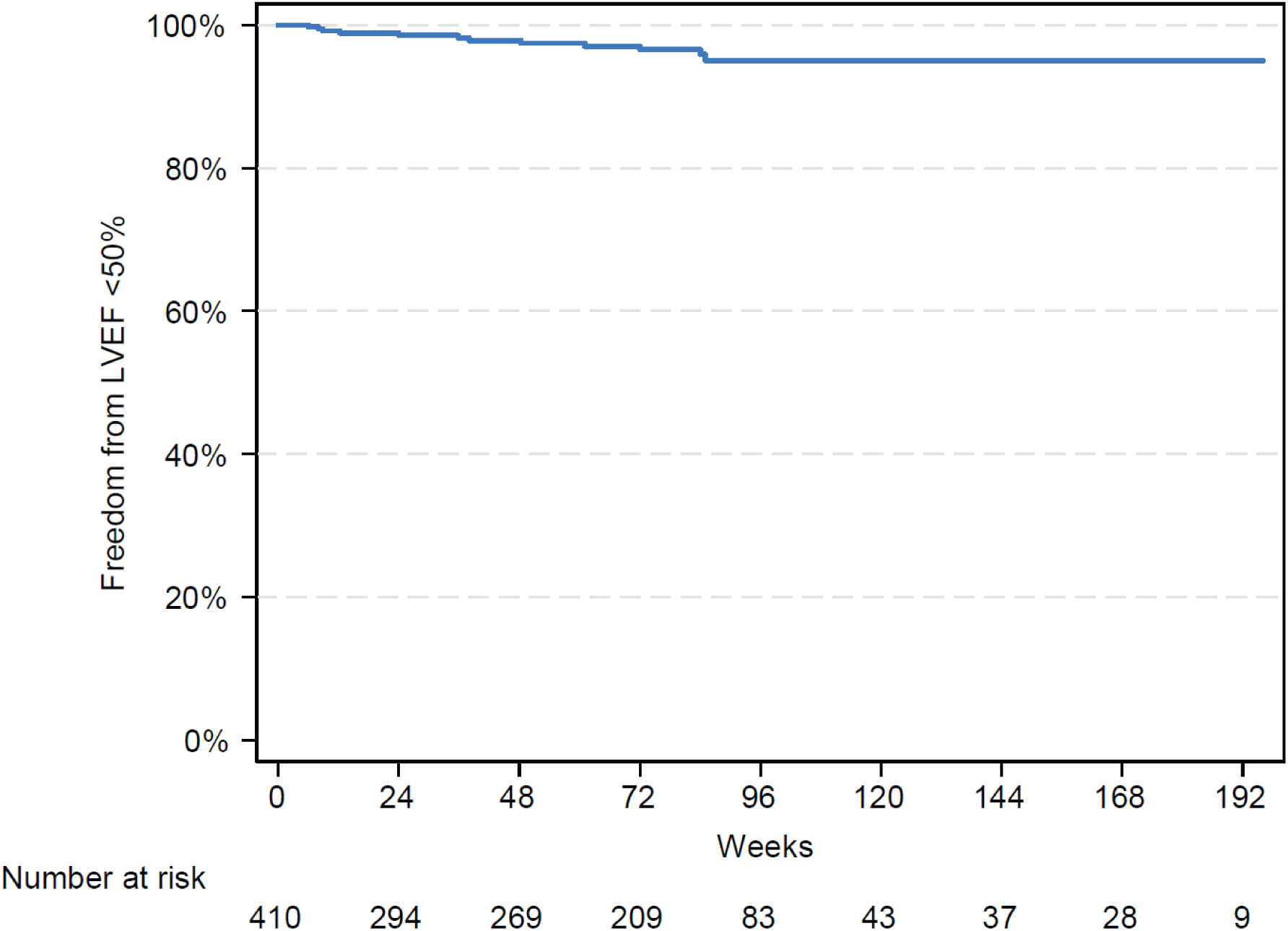
Freedom from LVSD defined as LVEF <50% Kaplan–Meier curve. LVEF, left ventricular ejection fraction; left ventricular systolic dysfunction.

**Table 2.** Patients with low EF events.

|  |  | Baseline |  |  |  |  | Event |  |  |  |  |  |  |  | Follow-up |  |  |
|---|---|---|---|---|---|---|---|---|---|---|---|---|---|---|---|---|---|
| Patient # | Age/ sex | Background therapy | LVEF (%) | Valsalva LVOT-G (mmHg) | NYHA Class | KCCQ-CSS | Lowest LVEF (%) | Core lab LVEF (%) | Week of lowest LVEF | Dose at lowest LVEF* (mg) | Valsalva LVOT-G (mmHg) | NYHA class | KCCQ-CSS | NT-proBNP % change from baseline | Next LVEF >50% | Days to LVEF >50% | Dose at next LVEF >50% (mg) |
| 1 | 41-45/M | BB | 69.4 | 59 | II | 97.92 | 47.3 | 52.86 | 9 | 15 | 7 | II | - | - | 60.2 | 58 | 10 |
| 2 | 56-60/M | CCB, DP | 67 | 74 | II | 78.13 | 49 | 51.18 | 48 | 20 | 16 | I | 97.92 | -90.10 | 55 | 13 | 15 |
| 3 | 71-75/F | BB | 65 | 61 | II | 73.44 | 45 | 54.74 | 72 | 15 | 4 | I | 94.79 | -83.80 | 55 | 7 | 10 |
| 4 | 56-60/M | BB | 61 | 84 | II | 90.1 | 47 | 54.72 | 61 | 20 | 31 | I | - | -69.60 | 54 | 14 | 15 |
| 5 | 51-55/M | BB | 70 | 79 | II | 71.88 | 45 | 66.55 | 85 | 15 | 4 | I | 100 | -94.80 | 55 | 7 | 10 |
| 6 | 56-60/M | BB, DP | 62 | 77 | II | 92.81 | 44 | 47.27 | 84 | 15 | 10 | II | 96.35 | -78.60 | 54 | 28 | 5 |
| 7 | 76-80/F | BB | 66 | 69 | III | 67.92 | 42 | 52.45 | 14 | 10 | 30 | III | 66.46 | -10.10 | 60 | 49 | 5 |
| 8 | 71-75/F | None | 55 | 42 | III | 28.13 | 42 | 54.88 | 20 | 5 | 53 | III | 28.13 | -35.60 | 52 | 14 | 5 |
| 9 | 41-45/M | BB, DP | 56 | 52 | II | 57.3 | 41 | 55.46 | 38 | 15 | 12 | II | 78.13 | -62.90 | 57 | 21 | 10 |
| 10 | 61-65/M | BB | 63 | 76 | II | 79.17 | 43 | 52.19 | 24 | 15 | 6 | II | 82.29 | -81.20 | 52 | 14 | 10 |
| 11 | 66-70/M | CCB | 65 | 86 | III | 25 | 44 | 54 | 24 | 15 | 12 | III | 34.38 | -24.00 | 55 | 14 | 15 |
| 12 | 71-75/F | BB | 66 | 90 | II | 95.83 | 49.9 | 58.8 | 36 | 20 | 8 | I | 96.35 | -90.30 | 57 | 14 | 15 |

\*Represents dose at the time of the lowest measured LVEF. No patients had an initial detection of LVSD while on 5mg.
BB, beta blocker; CCB, calcium channel blocker; DP, disopyramide; F, female; KCCQ-CSS, Kansas City Cardiomyopathy Questionnaire Clinical Summary Score; LVEF, left ventricular ejection fraction; LVOT-G, left ventricular outflow tract gradient; M, male; NYHA, New York Heart Association; NT-proBNP, N-terminal pro-B-type natriuretic peptide.

**Table 3.** Outcome Measures.

| **Site EF <50% (*N* = 12)** | **Baseline** | **At Occurrence<br>(*n* = 21)** | **Subsequent Follow-up** |
|---|---|---|---|
| LVEF site read, % | 63.8 (4.7) | 45.1 (2.53) | 51.7 (5.90) |
| LVEF core lab, % | 66.9 (8.43) | 55.4 (4.29) | 60.7 (7.94) |
| Resting LVOT-G site read, mmHg | 47.8 (34.06) | 8.7 (4.38) | 14.6 (9.13) |
| Valsalva LVOT-G site read, mmHg | 70.8 (14.58) | 27.6 (25.55) | 39.1 (23.26) |
| NYHA class, *n* (%) |  |  |  |
| I | 0 | 5 (25) | 6 (33.3) |
| II | 9 (75) | 8 (40) | 5 (27.78) |
| III | 3 (25) | 7 (35) | 7 (38.89) |
| IV | 0 | 0 | 0 |
| ≥1 NYHA class improvement, *n* (%) | - | 7 (35) | 7 (38.89) |
| NT-proBNP, median (IQR) | 716.5 (307.5–1200.0) | 131.5 (80.00–298.00) | 451 (125–596) |
| hs-cTn1, median (IQR) | 12.7 (8.2–17.0) | 9 (4.2, 10.4) | 14.6 (14.6–14.6) |

Subjects may have multiple low EF events. Data reported as mean (SD) or median (IQR 25%–75%) as appropriate for continuous data and *n* (%) for categorical data.
EF, ejection fraction; IQR, interquartile range; hs-cTn1, high-sensitivity cardiac troponin; LVEF, left ventricular ejection fraction; LVOT-G, left ventricular outflow tract gradient; NYHA, New York Heart Association; NT-proBNP, N-terminal pro-B-type natriuretic peptide; SD, standard deviation.

### Heart failure and other clinical events

Two patients developed non-serious signs or symptoms (dyspnoea and cardiac failure) of clinical heart failure at the time of echocardiographic LVSD, one of whom underwent a dose reduction followed by resolution of symptoms, improvement in LVEF and subsequently continued aficamten without further heart failure signs or symptoms. The second participant had a history of LVSD while randomized to placebo in the parent study (before enrolling in FOREST-HCM) and subsequently received aficamten in FOREST-HCM, which was permanently discontinued after developing LVSD while on drug. Among the 10 other patients who developed LVSD, all had resolution of LVSD with dose reduction. There were no deaths or heart failure hospitalizations in the reduced LVEF group. Two patients (0.5%) in the LVEF ≥50% group were hospitalized for heart failure, but both events were deemed unrelated to aficamten by investigators.

Among the 12 patients with LVSD, one participant experienced new-onset atrial fibrillation, and one participant experienced two recurrent atrial fibrillation events. All three atrial fibrillation events were temporally unrelated to the episodes of LVSD.

### Haemodynamic response

Aficamten therapy led to a substantial and sustained reduction in resting and Valsalva LVOT gradients (**Figure 3)**. The haemodynamic response to aficamten did not differ across systolic function subgroups. When LVSD did occur, the mean (SD) resting gradient was 8.7 ± 4.4 and the mean (SD) Valsalva gradient was 27.6 ± 25.6 mm Hg. At the subsequent visit, the resting gradient remained stable at 14.6 ± 9.1 mm Hg and the Valsalva gradient increased to 39.1 ± 23.3 mm Hg after the mandated dose reduction.

**Figure 3.**
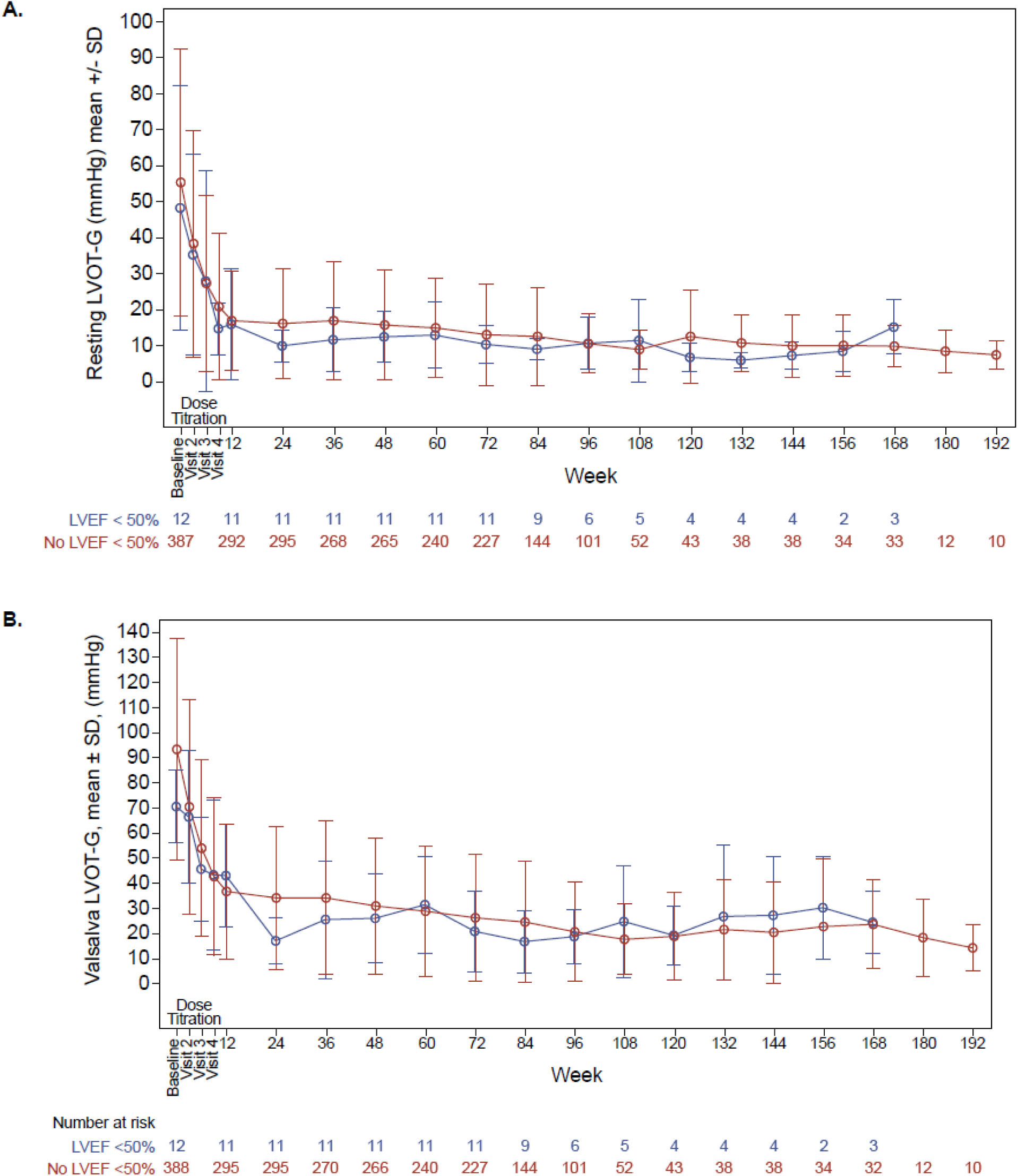
Resting (a) and Valsalva (b) LVOT gradients over time by subgroup. LVEF, left ventricular ejection fraction; LVOT, left ventricular outflow tract; SD, standard deviation.

### Cardiac biomarkers

Patients with LVSD did not demonstrate an increase in cardiac biomarkers at the time of the event. The median (25%, 75%) NT-proBNP level was 717 (308, 1200) ng/L at baseline and 132 (80, 298) ng/L at the time of LVSD. At the subsequent visit, the median (25%, 75%) NT-proBNP level increased to 451 (125, 596) ng/L after mandated dose reduction. Hs-cTnI levels followed a similar trajectory (**Table 3**).

*Subgroup analysis of patients with LVEF ≥50% to <55%*

A total of 47 patients (11.5%) had LVEF ≥50% to <55% at any time point (EAIR 8.8 per 100 patient-years) with 97 total occurrences representing 2.6% of all echocardiograms. The mean LVEF at the time of these events was 52.4 ±1.4% and no dose adjustments were made. At the subsequent visit, the mean (SD) LVEF improved to 56.5 ± 6.1%. Four patients (8.5%) experienced a subsequent LVEF <50% event (six events total), including the participant with systolic dysfunction while on placebo in the parent study. Haemodynamics remained stable, with mean (SD) resting and Valsalva gradients of 10.6 ±7.9 mm Hg and 24 ±19.5 mm Hg, respectively, at the time of LVEF ≥50 to <55%. At the subsequent visit, gradients were similar. NT-proBNP decreased from a median of 778 (349, 1604) ng/L at baseline to 125 (54, 257) ng/L at the time when LVEF ≥50% to <55%, and remained stable at 130 (64, 247) ng/L at the subsequent visit (**Supplemental Table 2**).

## Discussion

In this analysis from FOREST-HCM, treatment with aficamten guided by clinician-assessed echocardiographic monitoring was associated with a low incidence (2.9%) of LVSD (EAIR 2.1 per 100 patient-years), which was uniformly mild, and rarely associated with signs or symptoms of heart failure, with no serious events. Furthermore, dose reduction led to rapid recovery of LV function in nearly all cases, with the single outlier occurring in a participant with pre-existing systolic dysfunction while receiving placebo in the parent clinical trial. Importantly, there were no cases of persistent LVSD or LVEF <40% and no associated cases of serious heart failure, providing reassurance and underscoring the safety of aficamten. Notably, no patients receiving the 5 mg dose experienced a low LVEF event, supporting the inherent safety of lower-dose exposure.

A notable strength and source of novelty of the present analysis is that it reports outcomes according to the site-based echocardiographic assessments that drove treatment decisions. While core laboratory analyses remain important for standardization, clinicians prescribe, monitor, and adjust aficamten based on local echocardiographic interpretation. Therefore, these data provide a practical assessment of the safety and effectiveness of the aficamten monitoring and dose-adjustment paradigm as it is implemented in clinical practice.

The dose-titration and maintenance strategies implemented in FOREST-HCM, and now reflected in the product label, appear to be effective from a safety perspective. This is of great clinical importance from two perspectives: (1) LVSD with LVEF <50% appears to be well tolerated, without severe or irreversible clinical consequence, and can be managed safely with dose reduction; and (2) In the setting of an LVEF ≥50% to <55%, re-evaluation without adjusting the dose after 3 months is reasonable, since only a minority of patients subsequently developed LVSD. There appears to be no clinical consequences of an LVEF in this range, aside from a modestly elevated likelihood of detecting an LVEF <50% at a future visit. These findings provide important reassurance regarding the safety of aficamten when administered in accordance with protocol-guided echocardiographic surveillance and dose titration.

CMIs directly reduce excessive actin-myosin cross-bridge formation, and thereby lower contractile force, reduce systolic anterior mitral valve leaflet motion, and relieve dynamic LVOT obstruction. In patients with oHCM, baseline LVEF is often supranormal, reflecting hypercontractility. Consequently, modest reductions in LVEF are therapeutically desirable and reflect target engagement. This paradigm may extend to most patients who develop mild LVSD (LVEF ≥40% to <50%) while on aficamten. Specifically, the clinical and biomarker correlates of imaging-detected mild LVSD in the setting of aficamten are reassuring in that NT-proBNP levels did not increase in the presence of mild LVSD.

The thresholds for defining systolic dysfunction in the setting of aficamten treatment grew in a historical context based on clinical manifestation of patients with HCM who have extensive fibrosis and face progressive systolic dysfunction, ultimately leading to the possibility of heart transplantation.^13^ It is important to acknowledge that in this setting, while it is undesirable to breach this threshold, the clinical consequences appear to be very different. In fact, a key finding of this analysis is that patients appeared to derive meaningful clinical benefits from aficamten treatment even in the context of mild LVSD.

Approximately 12% of patients had at least one documented LVEF ≥50% and <55% following dose titration. However, their clinical outcomes including symptoms, haemodynamics, biomarkers, and adverse events were indistinguishable from patients who maintained LVEF ≥50% throughout treatment. These observations suggest that LVEF values in this range during aficamten treatment are clinically benign and do not need to be reflexively interpreted as treatment failure or toxicity.

Among the small subset of patients who developed LVSD, the majority quickly recovered LVEF following per protocol dose reduction. Notably, dose adjustment resulted in an increase in Valsalva LVOT gradients, albeit lower than at baseline. This underscores the need to carefully assess LVEF in this range to avoid unnecessary dose down-titration with potential detriments to clinical status. These data support the concept that clinicians should interpret LVEF in the context of patients’ symptoms, overall clinical trajectory, and cardiac biomarkers.

Heart failure signs or symptoms associated with reductions in LVEF were non-serious and rare, occurring in only 0.5% of treated patients. In one case, symptoms resolved and LVEF recovered following temporary interruption and dose adjustment, with no recurrent events after reinitiation at lower doses. The other participant who developed clinical heart failure in the context of LVSD had already presented with LVSD while on placebo in the parent aficamten trial, suggesting that this patient was developing progressive HCM-related LVSD unrelated to pharmacologic cardiac myosin inhibition. No deaths, cardiogenic shock, or progression to advanced heart failure therapies were observed. These findings suggest that the risk of clinically meaningful heart failure attributable to aficamten is low when patients are carefully selected and monitored. Importantly, the absence of severe systolic dysfunction in this cohort reinforces the effectiveness of the prespecified LVEF-guided titration strategy.

Experience with the first-in-class CMI mavacamten established the importance of echocardiographic monitoring and treatment interruption, guided to preserve systolic function owing to this agent’s pharmacodynamic properties. In the case of mavacamten, LVSD occurs modestly more frequently, and often in the context of additional factors contributing to reduced contractility or reflecting markers of more advanced myocardial disease, such as left bundle branch block, a history of atrial fibrillation or extensive late gadolinium enhancement.^4,14^ Despite the substantial size of the FOREST-HCM cohort, the rarity of LVSD events limits the ability to identify predictors of systolic dysfunction or develop a robust risk stratification model, highlighting the need for larger registries and longer-term follow-up. Notably, prolonged QRS duration was more common among patients experiencing low LVEF, suggesting that (similar to mavacamten) intraventricular conduction delays may influence susceptibility to LVSD with aficamten therapy. The shorter half-life and limited accumulation of aficamten contribute to the rapid recovery of systolic function observed after dose down-titration, largely without the need for treatment interruption. From a practical standpoint, these properties allow clinicians to respond more dynamically to echocardiographic findings, minimizing prolonged exposure to excessive contractile suppression.

The results of this analysis have direct implications for the real-world use of aficamten in oHCM. Routine echocardiographic surveillance remains essential. Mild reductions in LVEF can generally be managed conservatively with dose adjustment rather than discontinuation or interruption. Furthermore, LVEF values between ≥50% and <55% can safely be tolerated without dose reduction and do not appear to portend adverse outcomes in most patients. In addition, >90% of site-read instances of LVEF <50% had concurrent core laboratory interpretations of normal LVEF >50%, suggesting that echocardiographers at specialized HCM centres may prefer a more conservative interpretation erring on the side of safety in borderline cases.

### Limitations

Several limitations merit consideration. A strength of the analysis is the focus on site-read echocardiograms for dose adjustments, emulating real world clinical practice. This results in variable interpretation between site and core-read echocardiograms. In addition, the single-arm active treatment design without a placebo comparator limits definitively distinguishing treatment effects from background variation or other confounding factors. Furthermore, the small number of low LVEF events precluded formal multivariable analyses to identify independent risk factors for LVSD. In addition, NT-proBNP and KCCQ were not collected at every study visit in FOREST-HCM, limiting the ability to assess serial changes in biomarkers and patient-reported outcomes around the time of low LVEF events. Longer-term follow-up will be necessary to assess the durability of these findings and their applicability to broader patient populations.

## Conclusions

In this safety analysis from FOREST-HCM, aficamten was associated with infrequent, mild, and reversible reductions in LVEF, with a very low incidence of heart failure signs or symptoms and no cases of severe systolic dysfunction or serious heart failure. LVEF ≥50% to <55% was clinically benign. Together, these findings support the myocardial safety of aficamten and provide a practical framework for echocardiographic monitoring and dose optimization in patients with oHCM.

## Supporting information

IRB information

## Acknowledgements

The study team and the authors thank all patients who participated in this study and their families. They also thank all study sites and research personnel for their contributions. Editorial support was provided by David Sunter, PhD, on behalf of Engage Scientific Solutions, and was funded by Cytokinetics, Inc.

## Data availability statement

Qualified researchers may submit a request containing the research objectives, end points/outcomes of interest, statistical analysis plan, data requirements, publication plan, and qualifications of the researcher(s). In general, Cytokinetics does not grant external requests for individual patient data for the purpose of reevaluating safety and efficacy issues already addressed in the product labeling. Requests are reviewed by a committee of internal advisors, and if not approved, may be further reviewed by a Data Sharing Independent Review Panel.

Upon approval, information necessary to address the research question will be provided under the terms of a data sharing agreement. This may include anonymized individual patient data and/or available supporting documents, containing fragments of analysis code where provided in the analysis specifications. Requests may be submitted to.

## Abbreviations

CMI: cardiac myosin inhibitor
HCM: hypertrophic cardiomyopathy
hs-cTnI: high-sensitivity cardiac troponin I
KCCQ-CSS: Kansas City Cardiomyopathy Questionnaire–Clinical Summary Score
LVEF: left ventricular ejection fraction
LVOT: left ventricular outflow tract
LVSD: left ventricular systolic dysfunction
NT-proBNP: N-terminal pro–B-type natriuretic peptide
SRT: septal reduction therapy
TEAE: treatment-emergent adverse event

