## Supplemental Material for "Left ventricular systolic dysfunction in long-term aficamten therapy for obstructive HCM is rare, transient, and well-tolerated"

**Running title:** LVEF trajectory with aficamten in FOREST-HCM

**Supplemental Table 1.** Distribution of maintenance doses among groups. Data reported as N(%) and mean (standard deviation). a) 72 participants in the LVEF >=50% group had not yet reached maintenance dosing. b) p-value is from a two-sample t-test comparing maintenance dose between the LVEF<50% and LVEF>=50% groups.

| Maintenance Dose | Site-read LVEF<50% (N=12) | Site-read LVEF ≥50% (N=326)[a] | p-value[b] |
|---|---|---|---|
| 5mg | 2 (16.7) | 23 (7.1) | |
| 10mg | 2 (16.7) | 72 (22.1) | |
| 15mg | 7 (58.3) | 106 (32.5) | |
| 20mg | 1 (8.3) | 125 (38.3) | |
| Mean (SD) | 12.9 (4.5) | 15.1 (4.7) | 0.114 |

**Supplemental Table 2**. Outcome measures for occurrences of LVEF ≥50% and <55%. Subjects may have multiple events. Data reported as mean (SD) or median (IQR 25%–75%) as appropriate for continuous data and *n* (%) for categorical data.

EF, ejection fraction; hs-cTn1, high-sensitivity cardiac troponin; IQR, interquartile range; LVEF, left ventricular ejection fraction; LVOT-G, left ventricular outflow tract gradient; NYHA, New York Heart Association; NT-proBNP, N-terminal pro–B-type natriuretic peptide.

| **Site EF ≥50% to <55% (N=47)** | **Baseline** | **At Occurrence (N=97)** | **Subsequent Visit** |
|---|---|---|---|
| LVEF (%) site read | 64.5 (5.78) | 52.4 (1.37) | 56.5 (6.12) |
| LVEF (%) core lab | 68.7 (6.53) | 59.9 (7.33) | 60.8 (6.70) |
| Resting LVOT-G (mmHg) site read | 56.5 (45.94) | 10.6 (7.91) | 12.0 (9.07) |
| Valsalva LVOT-G (mmHg) site read | 84.6 (46.45) | 24.0 (19.45) | 27.6 (28.17) |
| NYHA class, n (%) | | | |
| I | 1 (2.1) | 57 (67.06) | 45 (61.64) |
| II | 29 (61.7) | 22 (25.88) | 24 (32.88) |
| III | 17 (36.2) | 6 (7.06) | 4 (5.48) |
| IV | 0 | 0 | 0 |
| ≥1 NYHA class improvement, n (%) | - | 62 (72.94) | 52 (71.23) |
| NT-proBNP, median (IQR) | 778 (349, 1604) | 124.5 (54, 257) | 130 (64, 247) |
| hs-cTn1, median (IQR) | 14.5 (9.1, 30.9) | 5.7 (3.6, 9.7) | 4.95 (3.5, 10.2) |

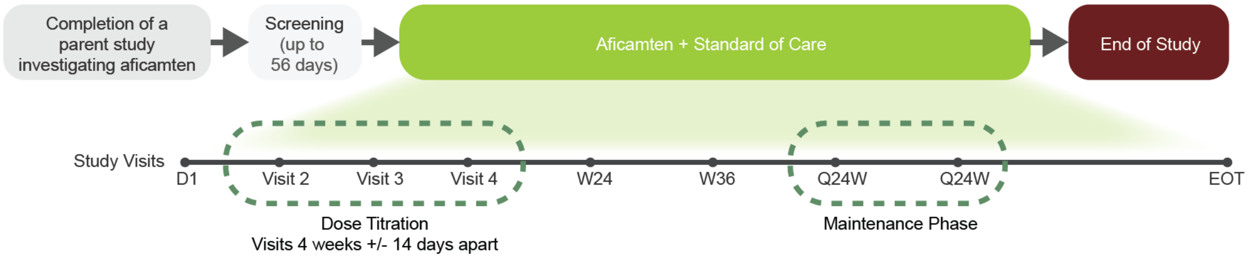


**Supplemental Figure 1**. FOREST-HCM Study Design
