## Supplementary material for "Left ventricular systolic dysfunction in long-term aficamten therapy for obstructive HCM is rare, transient, and well-tolerated": IRB information

The details of the IRB/oversight body that provided approval or exemption for the research described are given below:

The study was conducted in accordance with the Declaration of Helsinki and the International Council for Harmonisation Good Clinical Practice guidelines. The protocol was approved by the institutional review board or independent ethics committee at each participating site, and all participants provided written informed consent prior to enrolment: Comite de Etica en Investigacion ICBA, Comité Institucional de Ética de Investigación en Salud, Metro North Health HREC, Hospital das Clínicas - Hospital das Clínicas da Faculdade de Medicina da Universidade de São Paulo, Universidade São Francisco, Instituto Dante Pazzanese de Cardiologia, Investiga - Institutos de Pesquisa, Oncomed Centro de Prevenção e Tratamento de Doenças Neoplásicas, Nova Scotia Health Research Ethics Board, Comite d'ethique de le rescerche de l'institute Univeritaire de Cardiologie et de Pneumologie de Quebec - Univerite Laval, Research Ethics and New Technology Development Committee, Health Research Ethics Board, Biomedical Research Ethics Board (BREB) - University of Manitoba, Eticka komise Vseobecne fakultni nemocnice v Praze, The Medical Ethics Committees, Comité de Protection des Personnes Sud-Est IV, Ethik-Kommission der Universitaetsmedizin Goettingen, The National Ethics Committee, Medical Research Council, Ethics Committee for Clinical Pharmacology, The National Bioethics Committee, Sheba Helsinki Committee, Meir Helsinki Committee, Ziv Helsinki Committee, Hadassah Helsinki Committee, Comitato Etico Regione Toscana - Are Vasta Centro Azienda Ospedaliero - Univeritatia Carregi, [CEC] Comitato Etico Area Vasta Centro – Ufficio di Segreteria [LEC] Comitato Etico di AreaVasta Nord Oveset c/o Azienda OspedalieroUniversitaria Pisana, Comitato Etico Area Vasta Centro – Ufficio di Segreteria [LEC] Comitato Etico dell’Universita ‘Sapienza’ c/o Azienda OspedalieroUniversitaria Sant’Andrea, St. Luke's lnternational Hospital lnstitutional Review Board, Osaka General Medical Center Institutional Review Board, Osaka Medical and Pharmaceutical University Hospital Institutional Review Board, Osaka Metropolitan University Hospital Institutional Review Board, Kobe City Medical Center General Hospital Institutional Review Board, The Institutional Review Board, University of Tokyo Hospital, SHOWA Medical University Fujigaoka Hospital Institutional Review Board, Kochi Medical School Hospital Institutional Review Board, Hyogo Prefectural Harima-Himeji General Medical Center Institutional Review Board, Nippon Medical School Hospital Institutional Review Board, Medical Ethics Review Committee Erasmus MC, Komisja Bioetyczna Slaskiej Izby Lekarshkiej w Katowicach, CEIC – Parque da Saude de Lisboa, CElm-Parc de Salut Mar IMIM- Hospital del Mar Parc de Recerca Biomedica de Barcelona, South Central – Hampshire A Research Ethics Committee, Temple Quay House, WCG Institutional Review Board, Intermountain Institutional Review Board, Mayo Clinic Institutional Review Board, MUSC Office of Research Integrity, Vanderbilt University Medical Center Institutional Review Board, Northwestern University Institutional Review Board, Cleveland Clinic Institutional Review Board, University of Utah Institutional Review Board, Human Subjects Office IRB, Massachusetts General Brigham Institutional Review Board, University Hospitals IRB Administartion Office (Center for Clinical Research), UVA HSR-IRB, University of Minnesota, Human Research Protection Program (HRPP), Oregon Health & Science University Institutional Review Board, Houston Methodist Research Institute Institutional Review Board, New York Medical College Office of Research Administration, University of Pennsylvania Institutional Review Board, Advarra IRB, Cedars-Sinai Medical Center Institutional Review Board, IRB MED, Yale Human Research Investigation Committee, NYU Langone Health Institutional Review Board, Duke University Health System Institutional Review Board, Columbia University Medical Center Institutional Review Board
